# Leveraging paired serology to estimate the incidence of typhoidal *Salmonella* infection in the STRATAA study

**DOI:** 10.1101/2025.03.15.25324021

**Authors:** Jo Walker, Paula Russell, Leanne Kermack, Tan Trinh Van, Tran Vu Thieu Nga, Elli Mylona, Susana Camara, Young Chan Kim, Sonu Shrestha, Arne Gehlhaar, Josefin Bartholdson Scott, Farhana Khanam, Mila Shakya, Deus Thindwa, Melita A. Gordon, Buddha Basnyat, John D. Clemens, Firdausi Qadri, Robert S. Heyderman, Christine Dolecek, Susan Tonks, Thomas C. Darton, Andrew J. Pollard, Stephen Baker, James E. Meiring, Merryn Voysey, Virginia E. Pitzer, the STRATAA Study Group

## Abstract

Serologic surveillance of at-risk populations can be used to directly estimate the incidence of typhoidal *Salmonella* infection across a variety of settings, including those without access to facility-based blood-culture surveillance. We collected paired blood samples approximately three months apart from an age-stratified random sample of healthy children and adults in Bangladesh, Malawi, and Nepal as part of the Strategic Typhoid Alliance Across Asia and Africa (STRATAA) study. We used a multiplex bead assay to measure the concentration of IgG antibodies against seven *Salmonella* typhi/paratyphi antigens (CdtB, FliC, HlyE, LPSO2, LPSO9, Vi, and YncE) in each sample and identified recently infected participants by fitting a regression mixture model to the change in IgG concentration between participants’ samples. We estimated the seroincidence of infection in a Bayesian framework for each study site, age group, and antigen target. Finally, we compared the seroincidence estimates with crude and adjusted estimates of clinical incidence based on blood-culture surveillance. Seroincidence estimates were significantly higher than enteric fever incidence across all study sites, age groups, and antigen targets, even after adjusting for underreporting (median ratio: 25.4, interquartile range: 20.2-50.7). Seroincidence consistently peaked in the 0-4-year age group and declined moderately between children and adults (34% to 56% decline in HlyE seroincidence between the 5-9 and 30+ year old age groups), while enteric fever incidence peaked in older children and fell sharply in adults (71% to 95% decline in adjusted clinical incidence). Seroincidence estimates based on the HlyE and YncE antigens individually had the strongest correlation with observed enteric fever incidence across age groups and study sites (r = 0.63 and 0.71, respectively). These findings suggest that in endemic settings, both children and adults are frequently infected by typhoidal *Salmonella* serotypes, although only a fraction of these infections present as clinically identifiable enteric fever cases.

## Introduction

Enteric fever is an invasive bacterial disease caused by *Salmonella enterica* serotypes Typhi (*S.* Typhi) and Paratyphi A, B, or C (*S.* Paratyphi). Typhoidal *Salmonella* are spread through fecal-oral transmission, primarily via contaminated food and water(1). As a result, enteric fever mostly occurs in settings with limited clean water and sanitation in Africa and Asia, where it causes over 10 million cases and 100,000 deaths annually (2,3). While paratyphoid fever is rare in Africa, it often occurs in the same regions as typhoid fever in Asia, although usually at a lower incidence(3,4).

Existing diagnostics for enteric fever have several limitations affecting their utility in endemic settings(5,6). The Widal agglutination test detects antibodies against O and H antigens, and is simple and inexpensive to perform, but has relatively low sensitivity and specificity and requires paired samples for accurate interpretation(5,7). Blood culture is highly specific but only ∼60% sensitive, and requires several days to obtain a diagnosis(5,6,8). Bone marrow culture is ∼90% sensitive, but its invasiveness limits its scalability(5,8,9). Testing blood with a polymerase chain reaction (PCR) assay after a brief culture period offers improved sensitivity and speed over blood culture alone(5,9,10); however, both culture and PCR diagnostics require specialized training and equipment, which is not always available in endemic settings. Due to the lack of a cheap, accurate, and scalable diagnostic, routine enteric fever surveillance is rarely performed(2,3,11). The absence of surveillance data makes it difficult for policymakers to evaluate the local burden of disease and make evidence-based decisions about vaccines and other interventions(11).

Serosurveillance involves the collection and analysis of representative serologic data for the purpose of understanding population-level patterns of infection and immunity, and usually seeks to estimate either the incidence of infection (seroincidence) or the proportion of the population which has been infected (seroprevalence)(7,12). Serosurveillance has been used to characterize the frequency and natural history of infection, identify risk factors, evaluate the risk of future outbreaks, and guide public health policy for a variety of infectious diseases, including measles(13), rubella(14), polio(15,16), pertussis(17,18), COVID-19(19,20), and arboviruses(21,22).

Serologic studies of typhoidal *Salmonella* have most commonly used immunoglobulin G (IgG) against the Vi polysaccharide capsule produced by *S.* Typhi. However, anti-Vi IgG is often not elevated following infection, and the use of this antigen as the basis for typhoid conjugate vaccines (TCV) will confound its use as a marker of recent infection following vaccine introduction(7,23,24). Spurred by the need for improved diagnostics, high-throughput immunoscreening of enteric fever cases has identified additional antigen targets which may be promising indicators of infection(7,23,25–27). In particular, longitudinal studies following culture-confirmed enteric fever cases have shown that IgG antibodies against the Haemolysin E (HlyE) and lipopolysaccharide (LPS) antigens rise sharply and remain significantly elevated for months following *S.* Typhi and *S.* Paratyphi A infection(23,25,28,29). By modeling the kinetics of these antibodies and comparing them to cross-sectional measurements from the general population, *Aiemjoy et al*. have estimated seroincidence in multiple typhoid-endemic settings(7,25,30).

The Strategic Typhoid Alliance Across Asia and Africa (STRATAA) study was conducted at sites in Bangladesh, Malawi, and Nepal to improve our understanding of typhoid transmission and epidemiology across disparate endemic settings(4,31). Alongside enhanced clinical surveillance, a detailed household census, and healthcare utilization surveys, the STRATAA research consortium collected paired serological specimens from an age-stratified random sample of healthy participants at each study site. The incidence of seroconversion against the Vi antigen, defined as a two-fold rise in IgG on an ELISA assay, was higher than the adjusted incidence of typhoid fever and comparable between children and adults(4). In this study, we use a novel multiplex assay and a model-based cutoff approach to retest the STRATAA serologic samples, allowing us to precisely measure IgG and estimate seroincidence for seven typhoidal antigen targets.

## Methods

### Study design and enrollment of participants

The STRATAA study was performed in three urban communities: Mirpur thana in Dhaka, Bangladesh; Ndirande township in Blantyre, Malawi; and Lalitpur city in Kathmandu, Nepal. Among other factors, these areas differ in terms of demographics, population density, disease burden, and co-circulating pathogens; a detailed description of the sites is given in *Darton et al*(31). The baseline population of each site’s demarcated study area was enumerated in a household census conducted between June and October 2016(4). A final census was performed at each site two years later, with two intermediate census updates at the Bangladesh site and a single update at the Nepal site. At each site, the baseline census was used to randomly select participants for the serologic survey in five age groups: 0-4, 5-9, 10-14, 15-29, and 30+ years old. Each participant was randomly assigned to a 3-month window within the year for sampling to ensure representative coverage over the study period.

Participants were enrolled on an ongoing basis between February 2017 and February 2018 in Bangladesh, between December 2016 and April 2018 in Malawi, and between January 2017 and May 2018 in Nepal. However, almost no children at the Nepal site were enrolled in the serosurvey after November 2017, due to participation in the *TyVac-Nepal* TCV trial. Adults were not eligible for this trial and therefore continued to be enrolled in the serosurvey through the first half of 2018. To avoid confounding of age-specific incidence by calendar time, we excluded all participants at the Nepal site who were enrolled after November 2017.

### Sample collection and laboratory testing

Blood samples (1-3 ml) were collected in EDTA tubes from participants who could be located and provided informed consent. Where the randomized individual could not be identified, a household member in the same age group was selected where possible. Children younger than 6 months old who were identified in randomized households were also approached for enrollment to ensure this age group was present in the survey. All three sites performed anti-Vi IgG ELISAs using VaccZyme kits from The Binding Site Group (see *Meiring et al*. for results(4)). A further aliquot of blood was sent to either the University of Cambridge or Oxford University for standardized IgG testing with a custom multiplex bead-based immunoassay. Carboxylated fluorescent beads (Luminex-Diasorin) were coupled to purified S. Typhi antigens and incubated with the heat-inactivated sera at 1:20 dilution in assay diluent (phosphate-buffered saline, 0.05% Tween-20 and 1% BSA) for 1 hour in the dark on a shaker. The antigen panel included CdtB, FliC, HlyE, LPSO2, LPSO9, Vi, and YncE. IgG specific antibodies were detected with phycoerythrin (PE)-conjugated goat anti-human IgG (Moss.Inc) at 4µg/mL, incubated for 30 minutes in the dark on a shaker. Beads were washed and acquired on a Luminex 200 instrument. Results were collected as mean fluorescence intensity (FI) from duplicates.

### Data cleaning and processing

Per the study protocol, we planned to collect a baseline and follow-up sample from each participant at two visits approximately 90 days apart. In practice, some participants were lost to follow-up, while others had their second sample collected earlier or later than the 90-day post-baseline target. To minimize the impact of false-negative infections, which could occur if antibodies have enough time to undergo significant waning by the time the second sample is collected, we excluded participants with over 150 days between samples; we reduced this threshold to 100 days in a sensitivity analysis. We also excluded IgG measurements that had a non-positive FI value (indicative of measurement error), did not form a unique pair of baseline and follow-up measurements, or could not be linked to participant metadata.

To standardize measurements between antigen batches, we log10 transformed each FI value and calculated a z-score based on the batch-specific mean and standard deviation of the resulting log10(FI) values. We refer to the resulting output as the standardized fluorescence intensity (sFI).

### Classifying seroresponses

We fit a mixture of two linear regression models in which each qualifying participant is treated as a single observation, the sFI at visit 1 is the predictor variable, and the change in sFI between visits is the response variable. The fitted mixture model assigns each participant a posterior probability of membership in two clusters - one corresponding to a significant change (either increase or decrease) in anti-target-IgG between visits, and the other corresponding to stable IgG between visits. We fit the mixture model separately for each of the seven antigen targets and three study sites, for a total of 21 fitted models. We classified participants with a large rise in IgG between visits (posterior probability > 0.50) as infected, while all other participants were classified as uninfected. This process is illustrated in Figure 1 for the HlyE antigen target. As a sensitivity analysis, we also directly used each participant’s mixture-model-derived probability of infection to estimate seroincidence based on a modified likelihood equation, described below, rather than classifying participants as infected or not using the cutoff value of 0.5.

**Figure 1:**
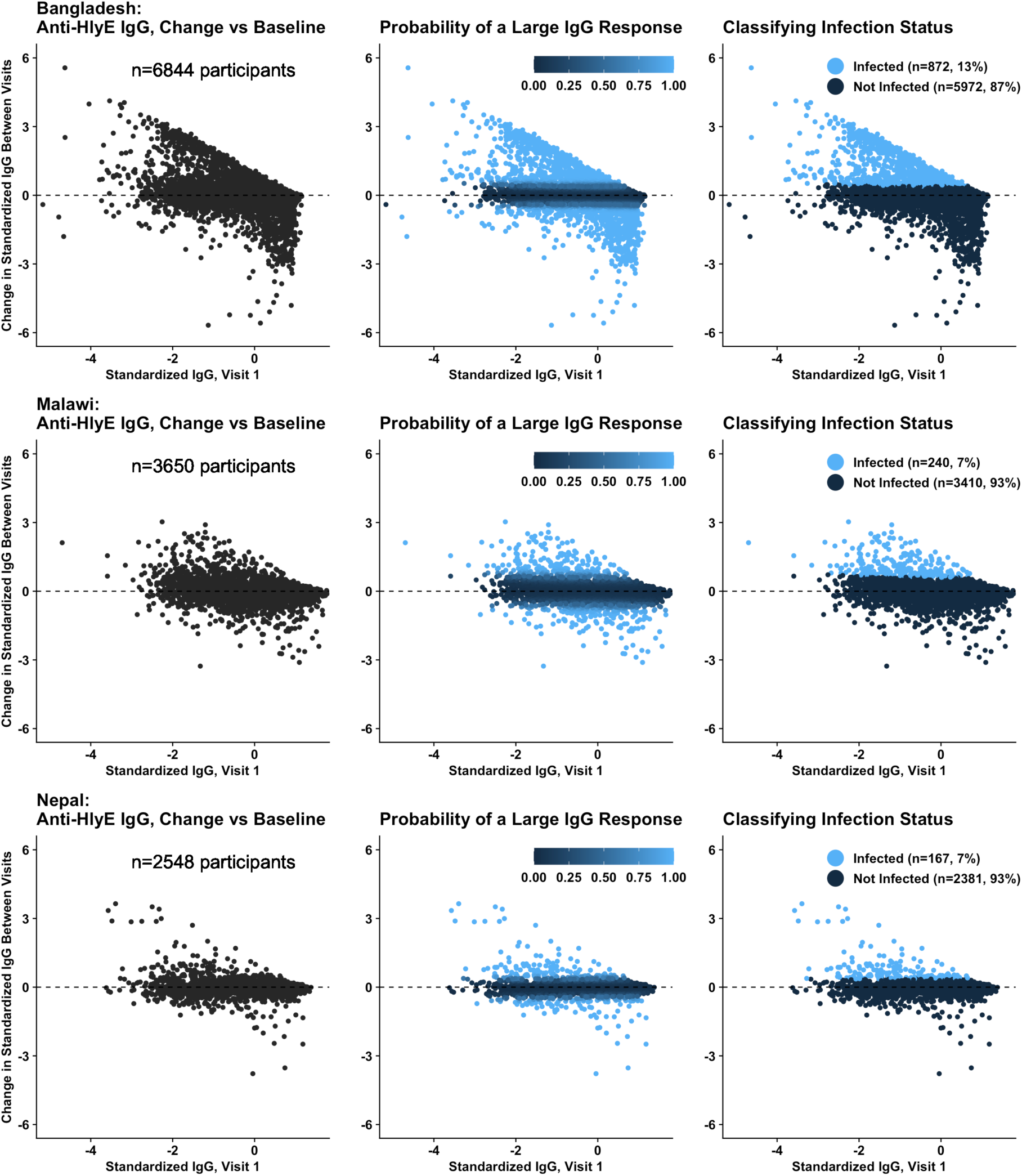
Classification of Anti-HlyE IgG Responses. IgG measurements refer to standardized log-transformed fluorescence intensity (see methods). Subfigures in the top, middle, and bottom rows correspond to the Bangladesh, Malawi, and Nepal study sites, respectively. *Left*: Each point represents the anti-HlyE IgG at baseline (X-axis) and the change in IgG from baseline to the follow-up visit (Y-axis) of a single participant. Most participants are clustered around the horizontal dashed line corresponding to no change in IgG between visits. *Middle*: Participants are colored by the posterior probability of having a large change (either an increase or decrease) in IgG between visits. This metric is derived from a two-cluster linear regression mixture model. *Right*: Participants who experienced a large rise in IgG between visits (posterior probability > 0.5) were classified as infected during this period. All other participants were considered to be uninfected.

### Estimating seroincidence

We assumed that participants experience new typhoidal *Salmonella* infections at a mean rate *λ* (seroincidence), such that the time *t* from visit 1 to the next infection is exponentially distributed. The likelihood function for *λ* is formed by linking infection status to the cumulative distribution function (CDF) of the exponential distribution:

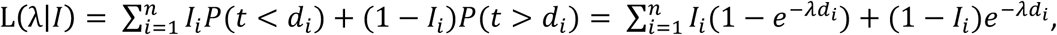

where *i* = 1,2, .. *n* denotes the set of participants, *I*_*i*_is a binary variable indicating whether or not participant *i* was infected between samples (*I*_*i*_= 1 or 0, respectively), and *d*_*i*_is the time interval between samples. We implemented this likelihood function in a Markov chain Monte Carlo (MCMC) framework with the Metropolis(32) algorithm to estimate the posterior distribution of *λ*, using an approximately flat gamma prior (shape = scale = 0.001), three independent chains of 10,000 samples, a 1,000 sample burn-in period, and a normal proposal distribution with a variance of 0.1 infections per person-year. We visually examined the output chains to confirm proper convergence. Seroincidence was estimated separately for each of the 3 study sites, 7 antigen targets, and 5 age groups. Finally, we calculated the “overall” age-standardized seroincidence for each study site and antigen as a census-weighted average of the age-specific posterior distributions of *λ*. As a sensitivity analysis, we replaced *I*_*i*_in the likelihood function with the posterior probability that participant *i* was infected, in order to account for uncertainty in the mixture model’s classification of infection status.

### Clinical surveillance

In order to evaluate the relationship between serologically-inferred infections and symptomatic disease at the population level, we compared our seroincidence estimates to the incidence of blood-culture-confirmed enteric fever (typhoid and paratyphoid fever) at each study site. In the STRATAA study, members of the household census population who presented to a study healthcare facility with either a recorded temperature of 38°C or a history of fever for ≥48 hours were approached for recruitment, and consenting participants provided blood for culture testing(4). At each study site, clinical surveillance activities and recruitment for the serologic survey began within a month of each other (Table 1). For Bangladesh and Malawi, we used the full ∼2-year clinical surveillance period to calculate enteric fever incidence, as this almost entirely overlaps with the interval covered by the serosurvey (Table 1). For Nepal, we only used the first year of clinical surveillance (2017) to calculate enteric fever incidence in order to align with the period covered by our serologic samples (see above). In addition to reported (unadjusted) incidence, we also present the symptomatic (adjusted) incidence of enteric fever, which has been corrected for blood culture sensitivity and the probability of care-seeking and testing(4).

**Table 1:** Participant Number and Characteristics.

|  | **Bangladesh** | **Malawi** | **Nepal** |
| --- | --- | --- | --- |
| **Serosurvey Enrollment Period** | Feb 2017-Feb 2018 | Dec 2016-Apr 2018 | Jan-Nov 2017 |
| **Clinical Surveillance Period** | Jan 2017-Dec 2018 | Nov 2016-Oct 2018 | Jan-Dec 2017 |
| **Total Participants** | 6,967 | 4,231 | 2,557 |
| **Excluded Participants:** | 115 | 578 | 8 |
| **Measurement error** | 11 | 0 | 0 |
| **No follow-up visit** | 39 | 38 | 1 |
| **Indeterminate visit date** | 3 | 12 | 3 |
| **>150 days between visits** | 62 | 528 | 4 |
| **Participants Included in Final Analysis** | 6,852 | 3,653 | 2,549 |
| **Participants by Age (%)** |  |  |  |
| 0-4 years | 1,914 (28%) | 704 (19%) | 197 (8%) |
| 5-9 years | 1,115 (16%) | 663 (18%) | 362 (14%) |
| 10-14 years | 702 (10%) | 417 (11%) | 314 (12%) |
| 15-29 years | 955 (13%) | 655 (18%) | 344 (13%) |
| 30+ years | 2,166 (31%) | 1,214 (33%) | 1,332 (52%) |
| **Days Between Baseline and Follow-Up Samples: Median (IQR)** | 84 (79 - 89) | 103 (91 - 113) | 110 (101-122) |

All modeling and analysis was performed in R (v3.6.3). The mixture model was fitted using the *flexmix* function of the *mixtools* R package (v1.2.0), which employs an expectation-maximization algorithm(33). Code and data is available at *github.com/pitzerlab/Typhoid-Seroincidence*.

### Ethics

For clinical surveillance and the serosurvey, Individual written informed consent was obtained from participants over the age of 18 or by a parent/guardian from participants below this age with additional assent sought from those aged between 11 and 17 years. For the healthcare utilization survey (used to adjust enteric fever incidence for underreporting) and the household demographic census, written informed consent was obtained from the head of each household (as the ‘key informant’) on behalf of the entire household. The STRATAA study protocol received ethics approval from the Oxford Tropical Research Ethics Committee (39–15), the Malawi National Health Sciences Research Committee (15/5/1599), the University of Malawi College of Medicine Research Ethics Committee, the Nepal Health Research Council (306/2015) and the icddr,b Institutional Review Board (PR-15199).

## Results

The number of participants meeting the inclusion criteria was 6,852 at the Bangladesh site, 3,653 at the Malawi site, and 2,549 at the Nepal site (Table 1). Follow-up samples were collected slightly earlier in Bangladesh than in Malawi and Nepal, with a median of 84, 103, and 110 days between visits, respectively. Baseline IgG levels against each target antigen were generally stable or slightly increased with age (particularly Vi) (Figure S1). Standardized IgG measurements for different antigens from the same participant sample were almost always positively correlated with each other, although most of the correlations were weak to moderate: Pearson’s correlation coefficients for antigen pairs ranged from −0.05 to 0.59 at the Bangladesh site, from 0.11 to 0.80 at the Malawi site, and from 0.17 to 0.75 at the Nepal site (Figure S2).

Seroincidence was considerably greater than enteric fever incidence across age groups, antigen targets, and study sites, even after adjusting for underreporting (Figure 2, median ratio: 25.4, IQR: 20.2-50.7). For instance, median age-standardized seroincidence estimates based on the HlyE antigen were 47.1 (95% CrI: 39.9 to 55.1) per 100 person-years (PY) in Bangladesh, 23.0 (95% CrI: 16.9 to 30.3) per 100 PY in Malawi, and 20.8 (95% CrI: 14.3 to 28.8) per 100 PY in Nepal (Table 2). These seroincidence estimates are 33, 49, and 18 times greater than the adjusted enteric fever incidence and 236, 361, and 260 times greater than the unadjusted enteric fever incidence at the Bangladesh, Malawi, and Nepal study sites, respectively.

**Figure 2:**
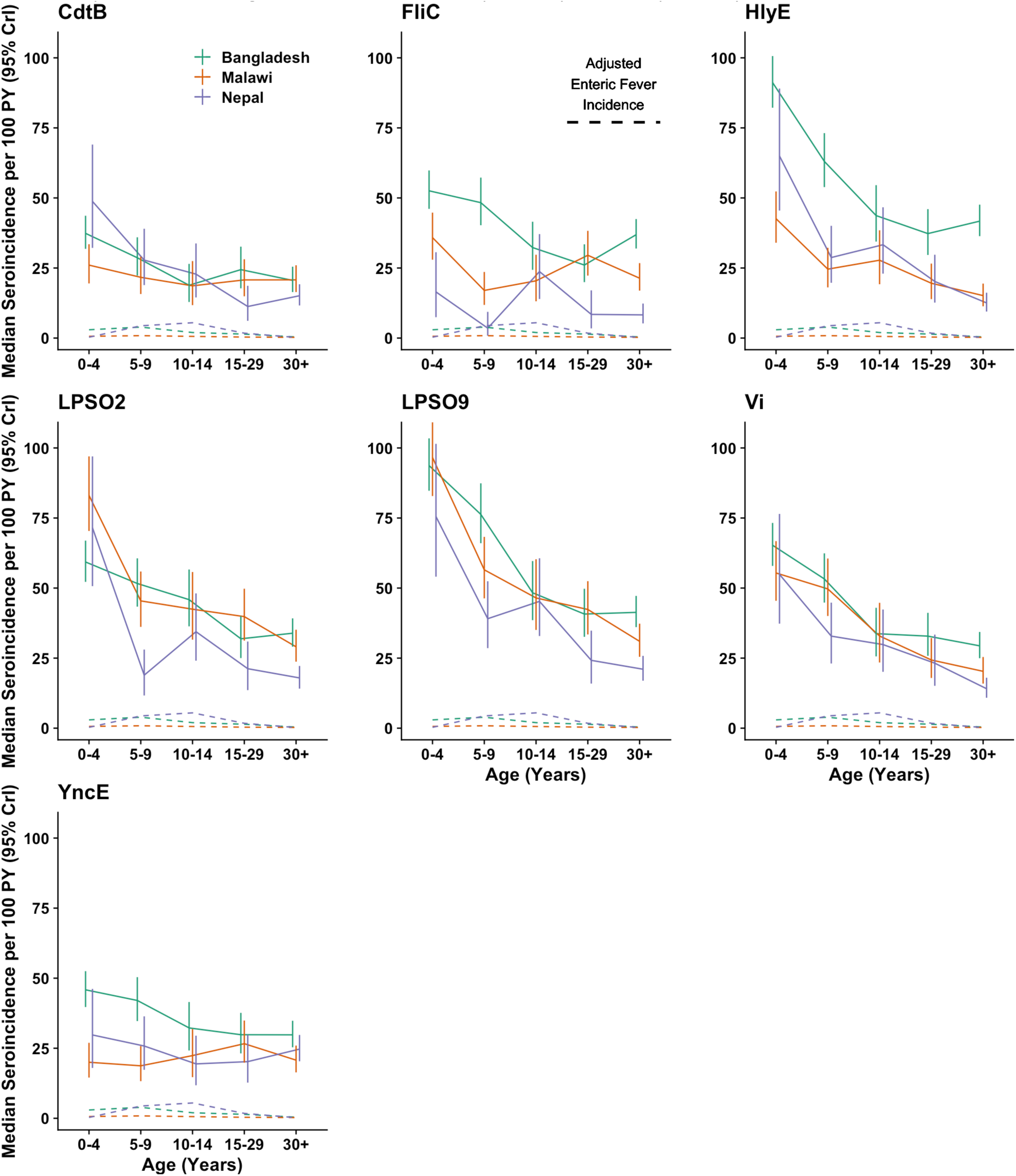
Seroincidence by Age, Antigen, and Study Site. Each subfigure corresponds to the specific antigen target which was used to classify participants’ infection status when calculating seroincidence. Solid lines denote the median seroincidence (y-axis) in each age group (x-axis). Vertical lines represent the 95% credible intervals of the seroincidence estimates, and dashed lines indicate the adjusted incidence of enteric fever. Green, orange, and purple lines correspond to the Bangladesh, Malawi, and Nepal study sites, respectively.

**Table 2:** HlyE Seroincidence and Clinical Enteric Fever Incidence by Age and Study Site.

| Age Group (Years) | Dhaka, Bangladesh |  |  |  | Blantyre, Malawi |  |  |  | Kathmandu, Nepal |  |  |  |
| --- | --- | --- | --- | --- | --- | --- | --- | --- | --- | --- | --- | --- |
|  | Unadjusted Enteric Fever Cases per 100 PY (95% CI) | Adjusted Enteric Fever Cases per 100 PY (95% CI) | Median HlyE Sero-incidence per 100 PY (95% CrI) | Ratio of Sero-incidence to Adjusted (Unadjusted) Enteric Fever Incidence | Unadjusted Enteric Fever Cases per 100 PY (95% CI) | Adjusted Enteric Fever Cases per 100 PY (95% CI) | Median HlyE Sero-incidence per 100 PY (95% CrI) | Ratio of Sero-incidence to Adjusted (Unadjusted) Enteric Fever Incidence | Unadjusted Enteric Fever Cases per 100 PY (95% CI) | Adjusted Enteric Fever Cases per 100 PY (95% CI) | Median HlyE Sero-incidence per 100 PY (95% CrI) | Ratio of Sero-incidence to Adjusted (Unadjusted) Enteric Fever Incidence |
| 0-4 | 0.47<br>(0.38-0.58) | 2.96<br>(1.99-4.79) | 91.17<br>(82.20-100.64) | 31<br>(194) | 0.08<br>(0.05-0.12) | 0.63<br>(0.40-0.96) | 41.66<br>(33.05-51.34) | 59<br>(467) | 0.03<br>(0.004-0.11) | 0.30<br>(0.04-1.20) | 64.96<br>(45.50-89.04) | 217<br>(2165) |
| 5-9 | 0.67<br>(0.55-0.81) | 3.91<br>(2.76-5.77) | 63.04<br>(53.87-73.14) | 16<br>(94) | 0.15<br>(0.10-0.20) | 0.86<br>(0.60-1.20) | 24.65<br>(18.19-32.49) | 30<br>(172) | 0.22<br>(0.11-0.39) | 4.38<br>(1.67-12.87) | 28.74<br>(19.72-40.06) | 7<br>(131) |
| 10-14 | 0.34<br>(0.26-0.44) | 1.98<br>(1.33-3.01) | 43.71<br>(34.42-54.57) | 22<br>(129) | 0.09<br>(0.06-0.13) | 0.60<br>(0.38-0.92) | 27.25<br>(18.74-37.99) | 38<br>(252) | 0.28<br>(0.17-0.43) | 5.47<br>(2.27-15.17) | 33.31<br>(22.98-46.67) | 6<br>(119) |
| 15-29 | 0.14<br>(0.11-0.18) | 1.41<br>(0.89-2.41) | 37.28<br>(29.65-46.02) | 26<br>(266) | 0.03<br>(0.02-0.05) | 0.36<br>(0.22-0.57) | 19.76<br>(13.96-26.89) | 57<br>(687) | 0.11<br>(0.08-0.16) | 1.75<br>(0.78-4.63) | 20.20<br>(12.65-29.74) | 12<br>(184) |
| 30+ | 0.04<br>(0.03-0.06) | 0.40<br>(0.23-0.74) | 41.72<br>(36.36-47.61) | 104<br>(1043) | 0.02<br>(0.01-0.04) | 0.25<br>(0.12-0.45) | 15.13<br>(11.35-19.57) | 52<br>(652) | 0.01<br>(0.005-0.03) | 0.22<br>(0.07-0.69) | 12.55<br>(9.46-16.18) | 57<br>(1255) |
| Overall | 0.20<br>(0.18-0.23) | 1.44<br>(1.14-1.48) | 47.11<br>(39.94-55.13) | 33<br>(236) | 0.06<br>(0.05-0.07) | 0.44<br>(0.35-0.72) | 22.96<br>(16.91-30.26) | 49<br>(361) | 0.08<br>(0.06-0.10) | 1.15<br>(0.71-1.98) | 20.79<br>(14.34-28.81) | 18<br>(260) |

Seroincidence was generally highest in the 0-4-year age group and declined gradually with age, but remained high even in adults (Figure 2). Exceptions include seroincidence based on the FliC antigen in Malawi and Nepal, and seroincidence based on the YncE antigen in Malawi, which were approximately stable with age. HlyE seroincidence was particularly high in the first two years of life at the Bangladesh and Malawi study sites, and among 2-3-year-olds at the Nepal site (Figure 3). The observed relationship between seroincidence and age contrasts with enteric fever incidence, which peaks in school-aged children before rapidly declining to a low level in adults (Figure 4). For instance, adjusted enteric fever incidence declined 90% at the Bangladesh site, 71% at the Malawi site, and 95% at the Nepal site between the 5-9 and 30+ year age groups, while HlyE seroincidence declined only 34%, 38%, and 56%, respectively, over the same interval.

**Figure 3:**
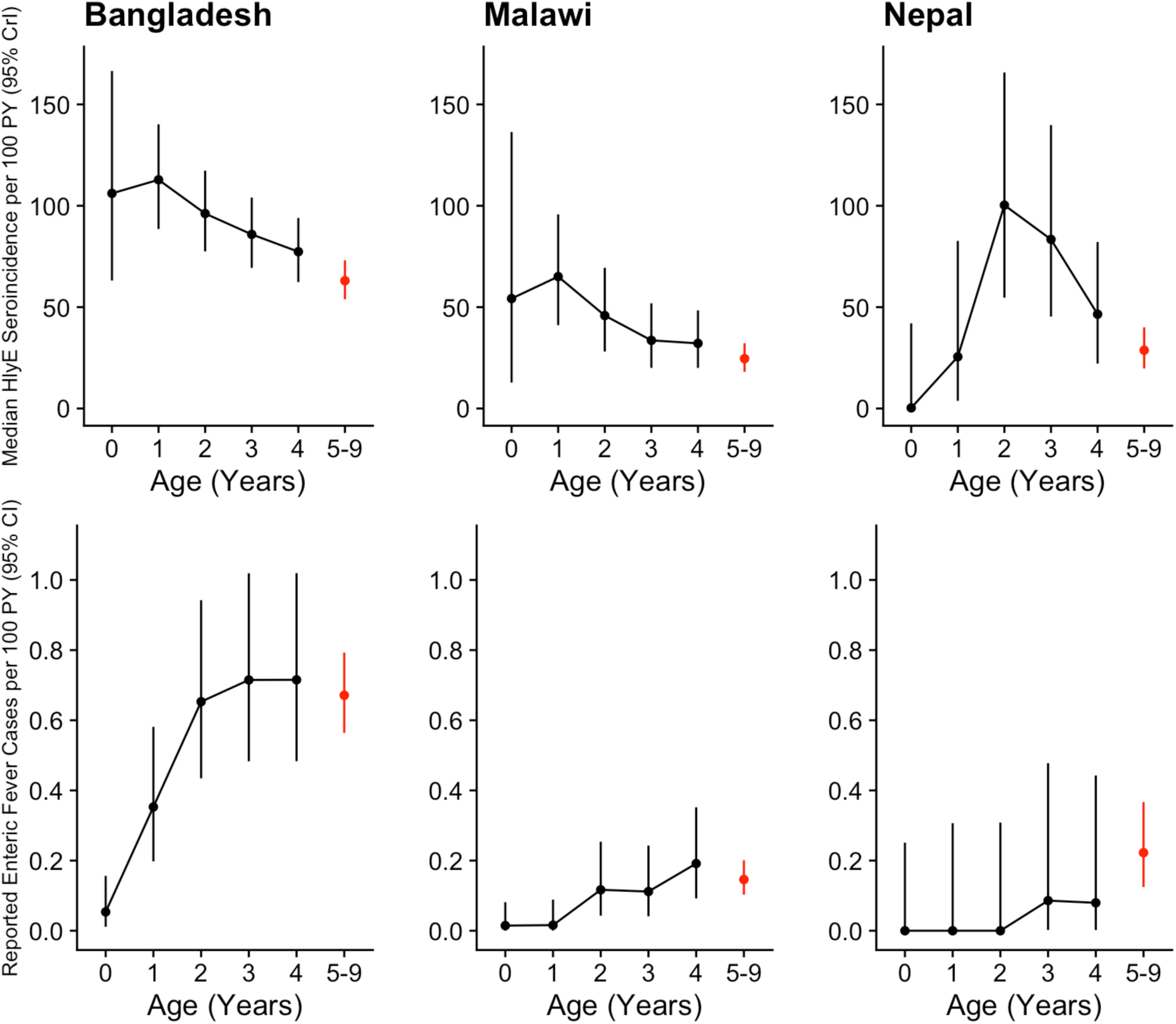
HlyE Seroincidence and Enteric Fever Incidence by Age in Young Children. Subfigures in the top and bottom rows display HlyE seroincidence and reported enteric fever incidence, respectively, at the Bangladesh (left), Malawi (middle), and Nepal (right) study sites. Age-specific incidence is shown for each of the first 5 years of life (black), and for 5-9 year-olds overall (red). Vertical lines represent 95% credible and confidence intervals for seroincidence and clinical incidence, respectively.

**Figure 4:**
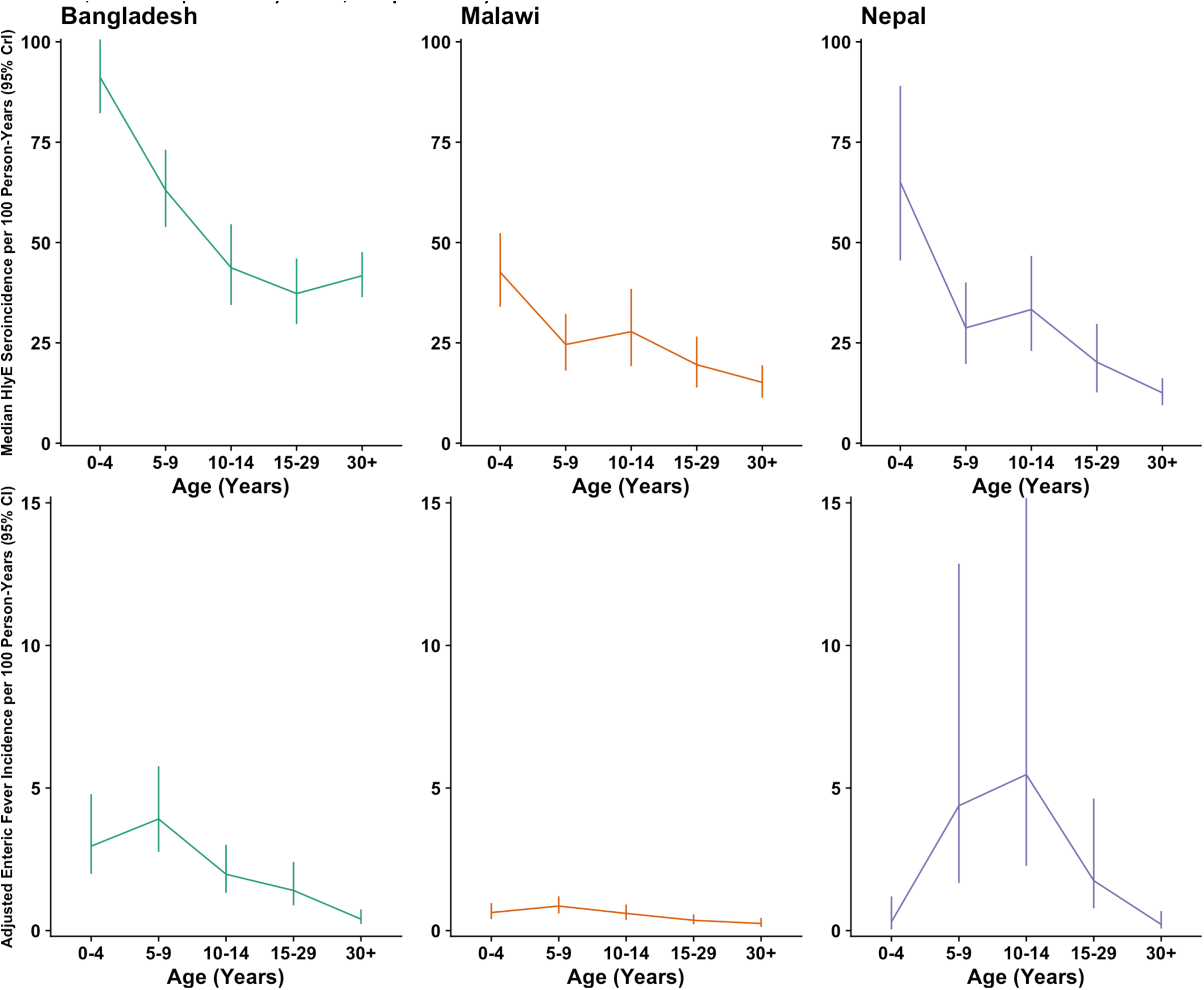
Relative Trend in HlyE Seroincidence and Enteric Fever Incidence by Age and Study Site. Each subfigure corresponds to a different study site. Solid lines represent the median seroincidence based on the HlyE antigen for each age group (left y-axis), and dashed lines represent the adjusted incidence of enteric fever cases (right y-axis). These measures are presented on different scales (compare axis labels) in order to highlight their association with age: seroincidence is highest in the 0-4-year age group and declines relatively gradually with age, while the incidence of enteric fever peaks in older children before rapidly declining to a low level in adults. Vertical lines represent the 95% credible intervals of seroincidence estimates. Green, orange, and purple lines correspond to the Bangladesh, Malawi, and Nepal study sites, respectively.

Seroincidence estimates were positively correlated with the reported incidence of enteric fever across different age groups and study sites (*r* = 0.19 to 0.71 across antigen targets) (Figure S3, Table 3). Adjusting the incidence of enteric fever for underreporting weakened its correlation with seroincidence (*r* = −0.14 to 0.29, Figure S4), although this appears to be attributable to the adjustment factors for Nepal: when estimates from the Nepal study site were excluded, the correlation between seroincidence and enteric fever incidence increased for all antigens, and was comparable between reported (*r* = 0.29 to 0.83) and adjusted enteric fever incidence (*r* = 0.27 to 0.87) (Table 3). For all comparisons, seroincidence estimates based on the HlyE and YncE antigens were most strongly correlated with enteric fever incidence, while seroincidence based on LPSO2 seroincidence exhibited the weakest correlation with enteric fever incidence. YncE was the only antigen target for which seroincidence captured the overall rank ordering of enteric fever incidence between study sites, with the highest incidence at the Bangladesh site, the lowest incidence at the Malawi site, and intermediate incidence at the Nepal site (Figure S5).

**Table 3:** Correlation Between Seroincidence and Enteric Fever Incidence Across Age Groups and Study Sites.

| Antigen Target | Correlation between seroincidence and reported enteric fever incidence |  | Correlation between seroincidence and adjusted enteric fever incidence |  |
| --- | --- | --- | --- | --- |
|  | Bangladesh, Malawi, and Nepal | Bangladesh and Malawi only | Bangladesh, Malawi, and Nepal | Bangladesh and Malawi only |
| CdtB | 0.24 | 0.64 | 0.13 | 0.68 |
| FliC | 0.60 | 0.73 | 0.13 | 0.74 |
| HlyE | 0.63 | 0.78 | 0.29 | 0.81 |
| LPSO2 | 0.19 | 0.29 | -0.14 | 0.27 |
| LPSO9 | 0.46 | 0.54 | 0.13 | 0.54 |
| Vi | 0.52 | 0.64 | 0.19 | 0.64 |
| YncE | 0.71 | 0.83 | 0.26 | 0.87 |

To evaluate temporal trends, we assigned each serosurvey participant to the month containing the midpoint between sample collection dates, and calculated HlyE seroincidence for each month and study site (Figure S6). At the Bangladesh site, seroincidence tracked the rise in enteric fever cases and blood-culture-positivity over the study period. In Malawi, both seroincidence and enteric fever cases peaked simultaneously at the beginning of 2017 and 2018, during the rainy season. At the Nepal study site, a peak in seroincidence in the spring 2017 coincided with low enteric fever cases, although this could be explained by the low number of patients enrolled in clinical surveillance in the early months of the STRATAA study(4). Subsequently, seroincidence tracked the decline in enteric fever cases and blood-culture positivity following the summer rainy season (June to August), but did not capture their resurgence in the last two months of 2017.

Excluding participants with over 100 days between samples did not have a large or consistent impact on seroincidence estimates (Figure S7). Seroincidence estimates were consistently lower (but still substantially higher than clinical enteric fever incidence) when we replaced binary infection status in the likelihood function with the posterior probability of infection from the mixture model (Figure S8).

## Discussion

In this study, we estimated the incidence of serologically-defined typhoidal *Salmonella* infection in three endemic urban settings by analyzing paired blood samples from the general population. These seroincidence estimates were substantially higher than the incidence of enteric fever across sites and age groups, even after adjusting for underreporting. This suggests that residents of endemic settings are infected by typhoidal *Salmonella* every few years on average, and that most of these infections are asymptomatic or do not present with traditional enteric fever symptoms. In addition to their difference in magnitude, seroincidence and enteric fever incidence followed different relative patterns with age: while seroincidence was highest in the first few years of life, enteric fever incidence was relatively low during this period and peaked later in childhood, as is typical in many typhoid-endemic settings(2,4,34–36). Our finding of higher seroincidence in young children is consistent with other serologic studies(4,25), and contradicts the idea that enteric fever incidence is low in young children because they are not exposed as often as older children. Cases in young children may have been under-detected in the STRATAA study, relative to other age groups(37). There is some evidence that the presentation of enteric fever is less specific, and potentially less severe, in young children, which could affect care-seeking, and parents of young children may have been less likely to consent to the collection of blood for culture diagnosis(1,38,39). Blood samples from young children, particularly infants, are also more difficult to collect and lower in volume than those from older patients, reducing culture sensitivity(8). While the adjustment factors applied to the culture-confirmed incidence data in the STRATAA study varied by age and attempted to control for some of these factors, the methodology may not have fully captured all of the differences between age groups. The low incidence of enteric fever in adults, on the other hand, likely reflects the gradual development of adaptive immunity from repeated exposure events throughout the lifespan(40). Our seroincidence results reveal that the low burden of disease in adults only can be partially explained by a reduced risk of infection, which suggest that infections in this group are also less likely to result in symptomatic disease. The relatively high seroincidence in adults suggests that they are frequently infected by typhoidal *Salmonella*, and therefore may be more important drivers of transmission than their disease burden alone would suggest; this may explain why only vaccinating children <15 years old failed to generate strong indirect protection in a cluster-randomized trial of TCV in Dhaka(41).

Seroincidence estimates based on the HlyE and YncE antigen targets were most strongly correlated with enteric fever incidence between age groups and settings. In a comparison of longitudinal antibody responses in culture-diagnosed enteric fever cases, a previous study showed that anti-HlyE IgG levels increased sharply following diagnosis and remained significantly elevated, relative to healthy controls from the community and culture-negative patients, for at least three months in both typhoid and paratyphoid A cases(23). Anti-YncE IgG levels, which have primarily been studied as a potential marker of chronic *S.* Typhi carriage(42), also rose and remained elevated following the onset of enteric fever, but these responses were much less distinctive against the background level of antibodies in non-cases. Due to their strong association with enteric fever cases at both the individual and population level, antibody responses against the HlyE antigen should be prioritized as an endpoint in future seroepidemiology studies.

Our seroincidence estimates were similar in magnitude to those obtained with an alternative approach leveraging cross-sectional serology data(25). In both studies, overall seroincidence was >5 per 100 person-years for each study site and antigen, and were substantially higher than the adjusted incidence of clinical disease in their respective populations. Both studies included estimates from sites in Dhaka and Kathmandu, and the ratio of overall seroincidence to adjusted clinical incidence was similar between the two studies in these locations (33 vs 32 in Dhaka, and 18 vs 16 in Kathmandu), which suggests that both methods produce similar results. Since paired and cross-sectional samples resulted in similar seroincidence estimates, the logistical benefit of not having to collect a follow-up sample may make cross-sectional sampling the more attractive option for future seroepidemiology studies of typhoidal *Salmonella*. By contrast, our Vi seroincidence estimates were much higher than the previously-published ELISA-based Vi seroincidence estimates from the STRATAA study, despite being based on the same target antigen and participant samples (Figure S9). The multiplex bead assay used in this study may have been able to measure IgG antibodies more precisely than the traditional ELISA method, which would have allowed us to detect serological exposures more easily.

Our analysis is subject to several limitations which should be considered when interpreting our results. First, we assumed that a large rise in IgG against the target antigen indicated typhoidal *Salmonella* infection, but this response could also arise from cross-reactive exposures to related bacteria. Cross-reactivity would inflate our seroincidence estimates and weaken their association with clinical enteric fever cases at the population level. As described above, anti-HlyE IgG has a strong and specific association with recent enteric fever cases(23,25); as a result, seroincidence estimates based on this target should be fairly robust to the impact of cross-reactive exposures. Second, our approach is premised on the idea that IgG will rise sharply and remain significantly elevated for several months following infection. This was demonstrated previously in a longitudinal study of typhoid and paratyphoid A fever cases, but it is possible that the magnitude and shape of antibody responses varies between clinical and asymptomatic cases(23). If asymptomatic infections only result in a small or transient rise in IgG antibodies, then some participants could have been misclassified as uninfected. Third, our methodology assumes that seroincidence is constant over time. In reality, incidence varies seasonally and from year-to-year, and therefore our estimates should be treated as averages over the study period. Incidence can also vary between subgroups of participants with different risk factors.

We attempted to control for this by recruiting participants from a randomly selected sample of residents enumerated through the STRATAA household census. Finally, with the exception of Vi (which is only present in *S.* Typhi), the target antigens in this study are expressed by both *S.* Typhi and *S.* Paratyphi. As a result, seroincidence estimates based on these antigens do not distinguish between the two serotypes. However *S.* Paratyphi is not believed to circulate at the Malawi study site, so exposures in this setting can be attributed to *S.* Typhi(4).

We conducted a seroepidemiologic study of typhoidal *Salmonella* infection dynamics at three study sites in Bangladesh, Malawi, and Nepal. We found that the seroincidence of infection was much higher than symptomatic enteric fever incidence, varied with age, and was predictive of the population-level disease burden, particularly for the HlyE and YncE antigen targets. These findings strengthen our understanding of the transmission dynamics and natural history of *S.* Typhi and *S.* Paratyphi serotypes, and can inform the design of future serologic studies seeking to generate data on these pathogens.

## Data Availability

Deidentified data is available online at github.com/pitzerlab/Typhoid-Seroincidence

## Acknowledgements

We acknowledge the contributions of the participants who took part in the STRATAA study and the large field and laboratory teams at the three study sites, including Amit Aryja, Binod Lal Bajracharya, David Banda, Yama Mujadidi, Pallavi Gurung, Arifuzzaman Khan, Clemens Masesa, Tikhala Makhaza Jere, Archana Maharjan, George Mangulenji, Maurice Mbewe, Harrison Msuku, Nirod Chandra Saha, Prasanta Kumar Biswas, Anup Adhikari, and the Nepal Family Development Foundation team.

---

**Supplementary Figure 1:**
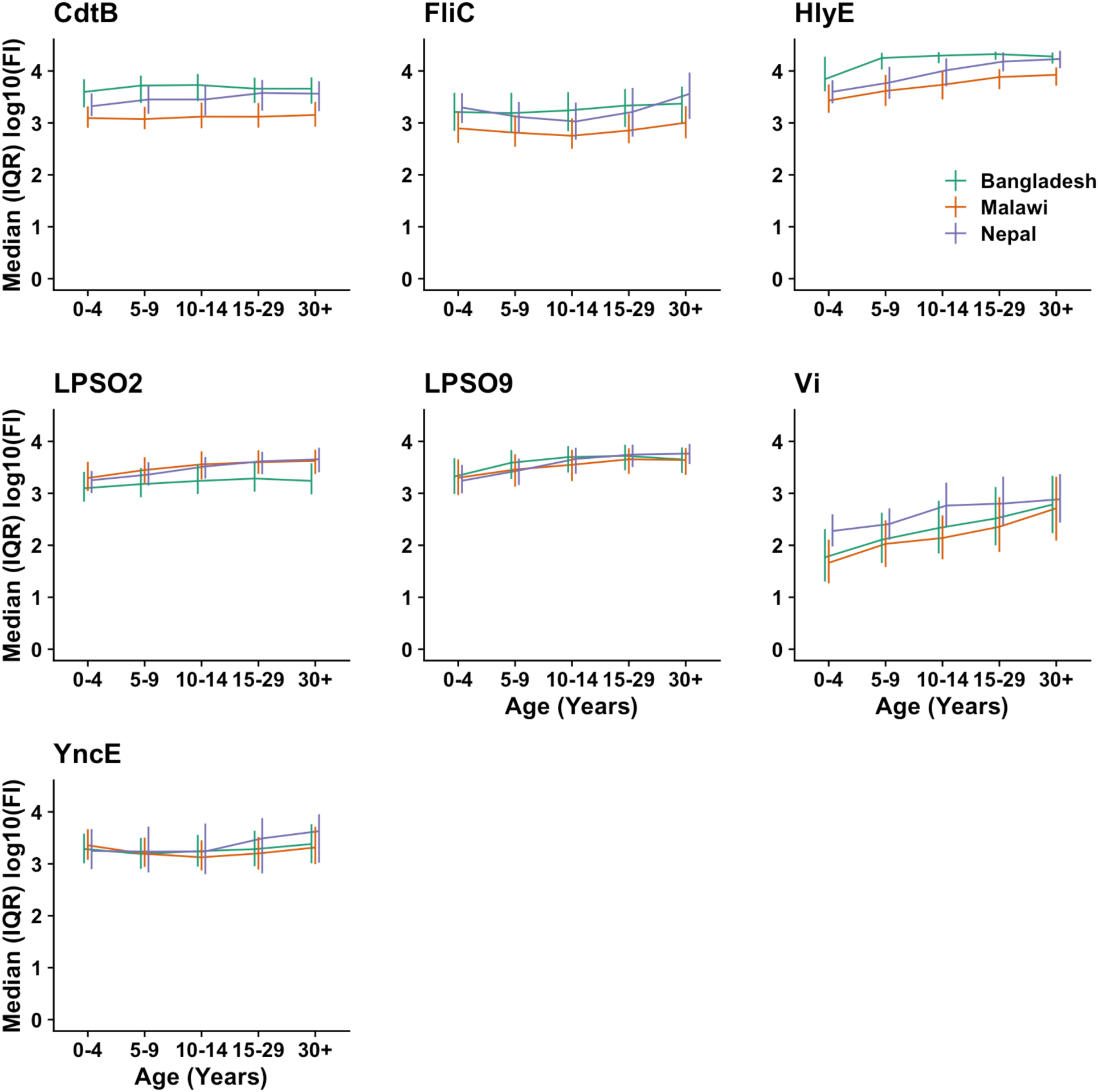
Baseline IgG by Age and Study Site. Each subfigure corresponds to a specific antigen target against which IgG antibodies were measured. Solid lines denote the median of the log10-transformed fluorescence intensity (FI, a proxy for IgG concentration, y-axis) across participant’s baseline samples in each age group (x-axis). Vertical lines represent the interquartile range of the log10(FI) measurements. Green, orange, and purple lines correspond to the Bangladesh, Malawi, and Nepal study sites, respectively.

**Supplementary Figure 2:**
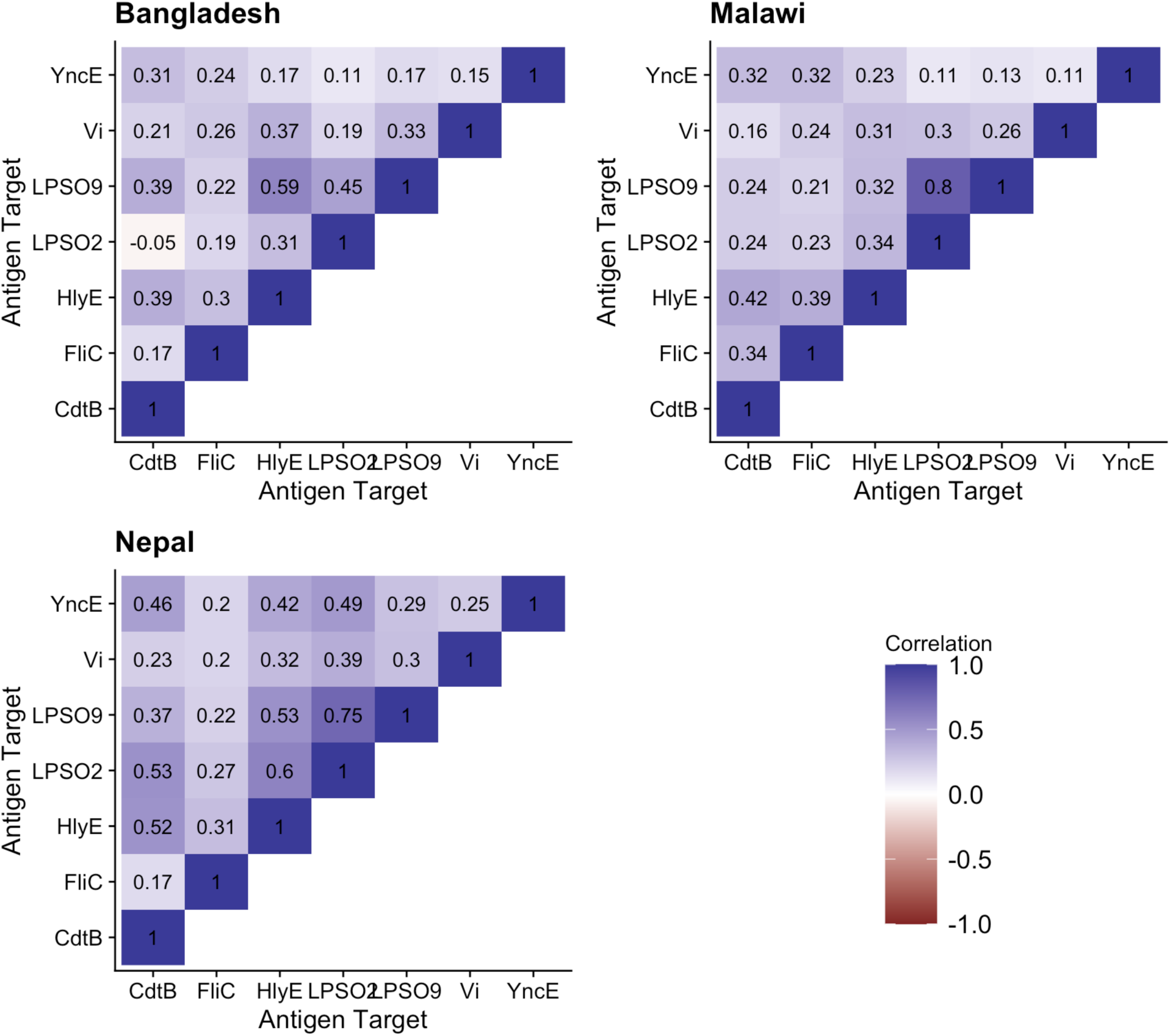
Correlation of IgG Between Antigen Targets. This figure displays the Pearson’s correlation coefficient of standardized IgG measurements from the same participant sample for each unique pair of antigen targets.

**Supplementary Figure 3:**
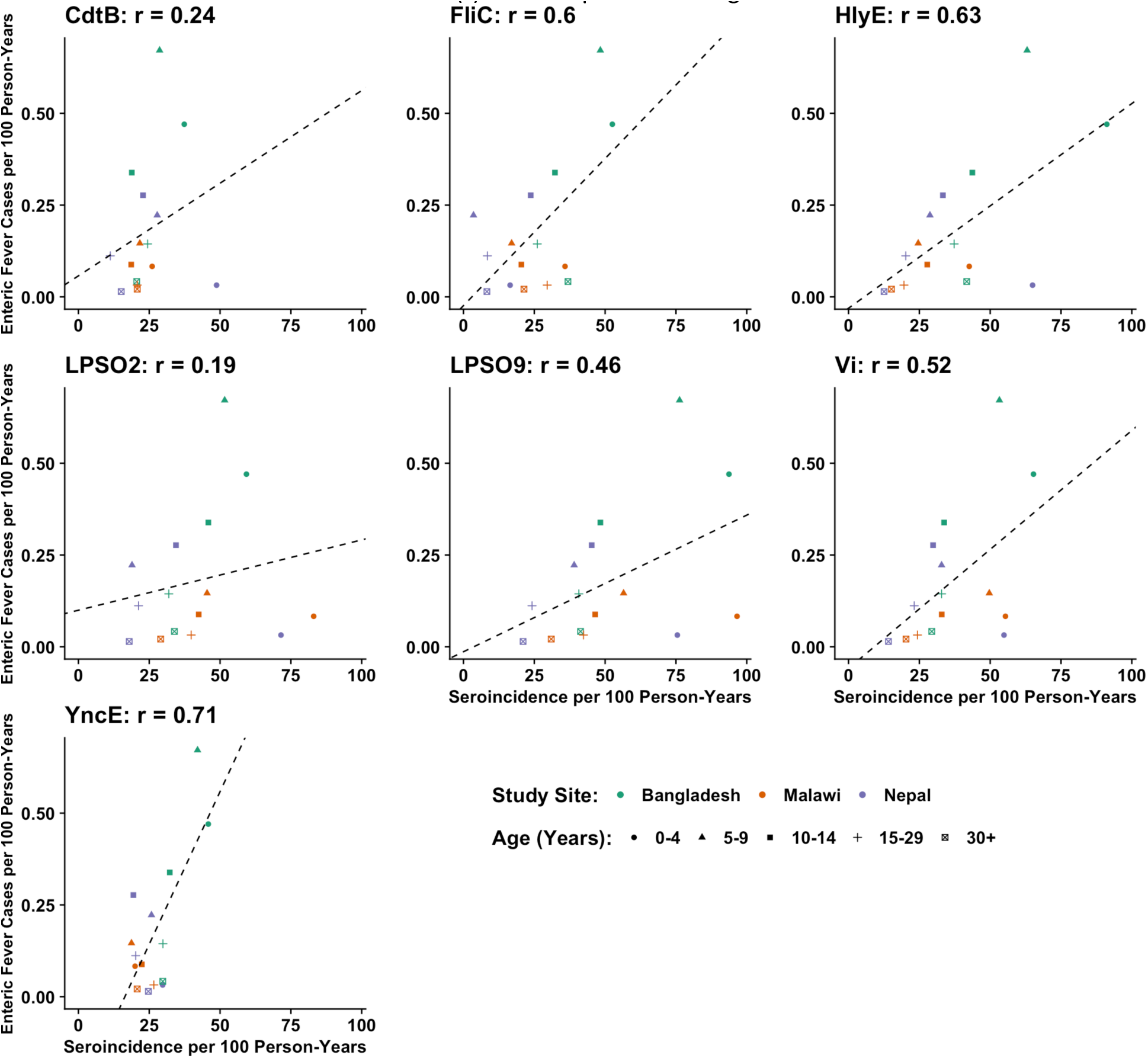
Correlation Between Seroincidence and Reported Enteric Fever Incidence. Each subfigure corresponds to the specific antigen target which was used to classify participants’ infection status when calculating seroincidence. Each point corresponds to an age group (shape) at a given study site (color). The position of each point represents the seroincidence (x-axis) and unadjusted enteric fever incidence (y-axis) for that age group and study site during the study period. The linear relationship between seroincidence and enteric fever incidence is indicated by a dashed line of best fit and the Pearson’s correlation coefficient (r) at the top of the subfigure.

**Supplementary Figure 4:**
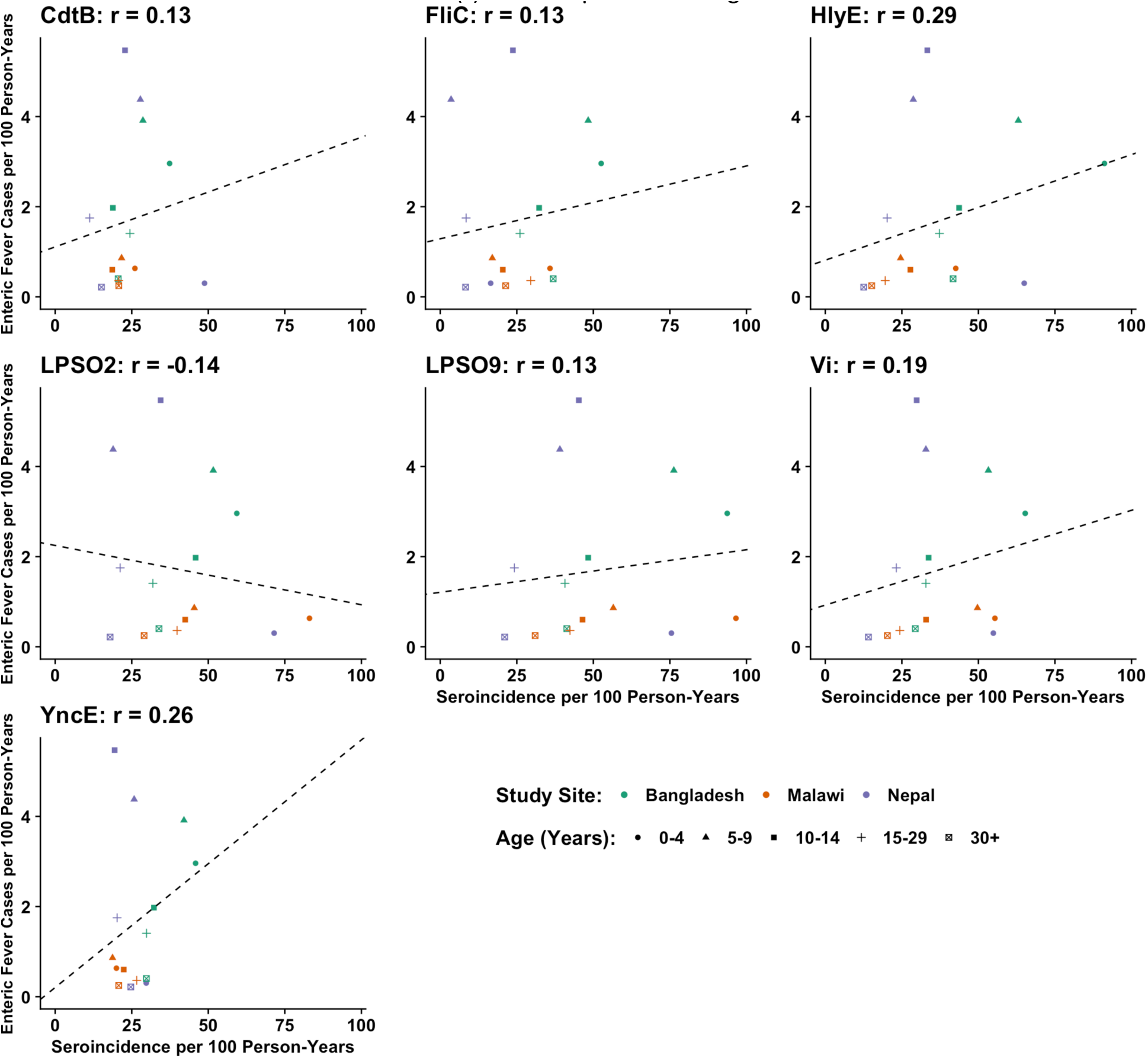
Correlation Between Seroincidence and Adjusted Enteric Fever Incidence. Each subfigure corresponds to the specific antigen target which was used to classify participants’ infection status when calculating seroincidence. Each point corresponds to an age group (shape) at a given study site (color). The position of each point represents the seroincidence (x-axis) and adjusted enteric fever incidence (y-axis) for that age group and study site during the study period. The linear relationship between seroincidence and enteric fever incidence is indicated by a dashed line of best fit and the Pearson’s correlation coefficient (r) at the top of the subfigure.

**Supplementary Figure 5:**
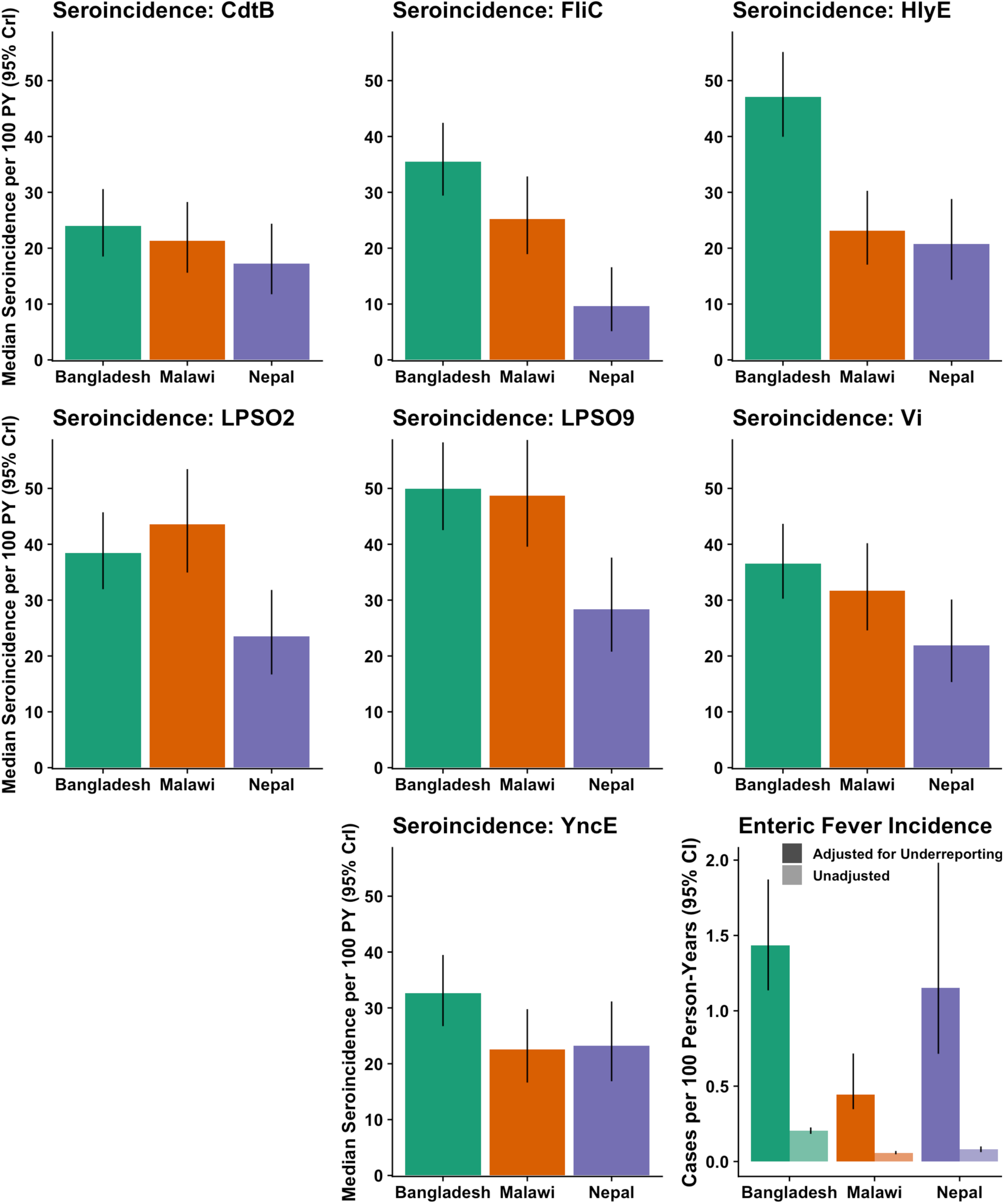
Overall Age-Standardized Seroincidence and Enteric Fever Incidence by Study Site. Each bar corresponds to a different study site, and the bar heights represent either the seroincidence of infection or the incidence of enteric fever cases at each site, as indicated. Incidence measures in this figure correspond to the overall population at each study site and its unique age distribution, rather than any specific age stratum.

**Supplementary Figure 6:**
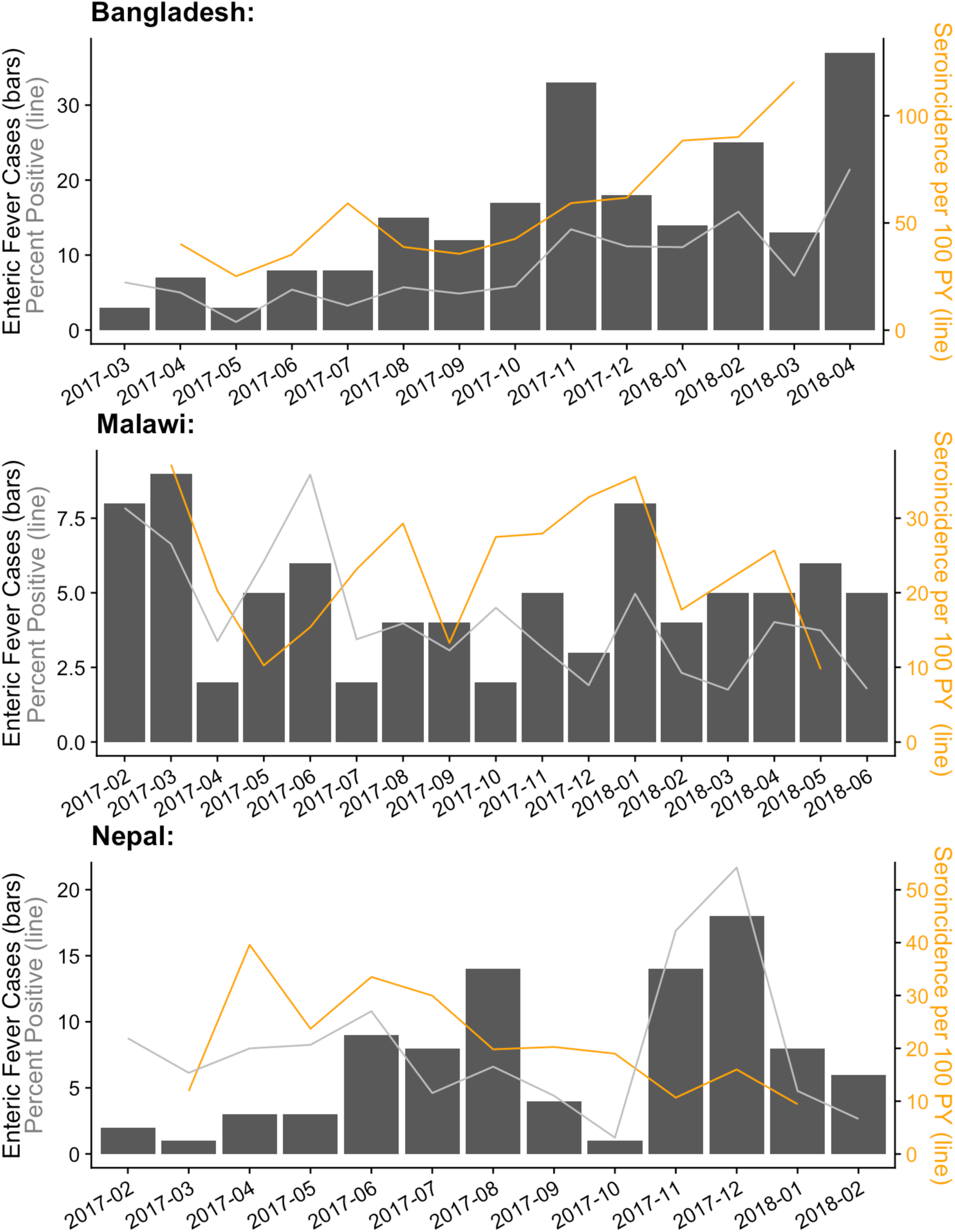
Monthly Enteric Fever Cases and HlyE Seroincidence. The height of each bar indicates the monthly number of blood-culture-confirmed enteric fever cases. The orange line represents the monthly estimated HlyE seroincidence, with each participant assigned to the month of the midpoint between their baseline and follow-up samples. The top, middle, and bottom subfigures correspond to the Bangladesh, Malawi, and Nepal STRATAA sites, respectively. Seroincidence estimates based on fewer than 50 participants are suppressed (April and June 2018 in Bangladesh and Malawi, respectively).

**Supplementary Figure 7:**
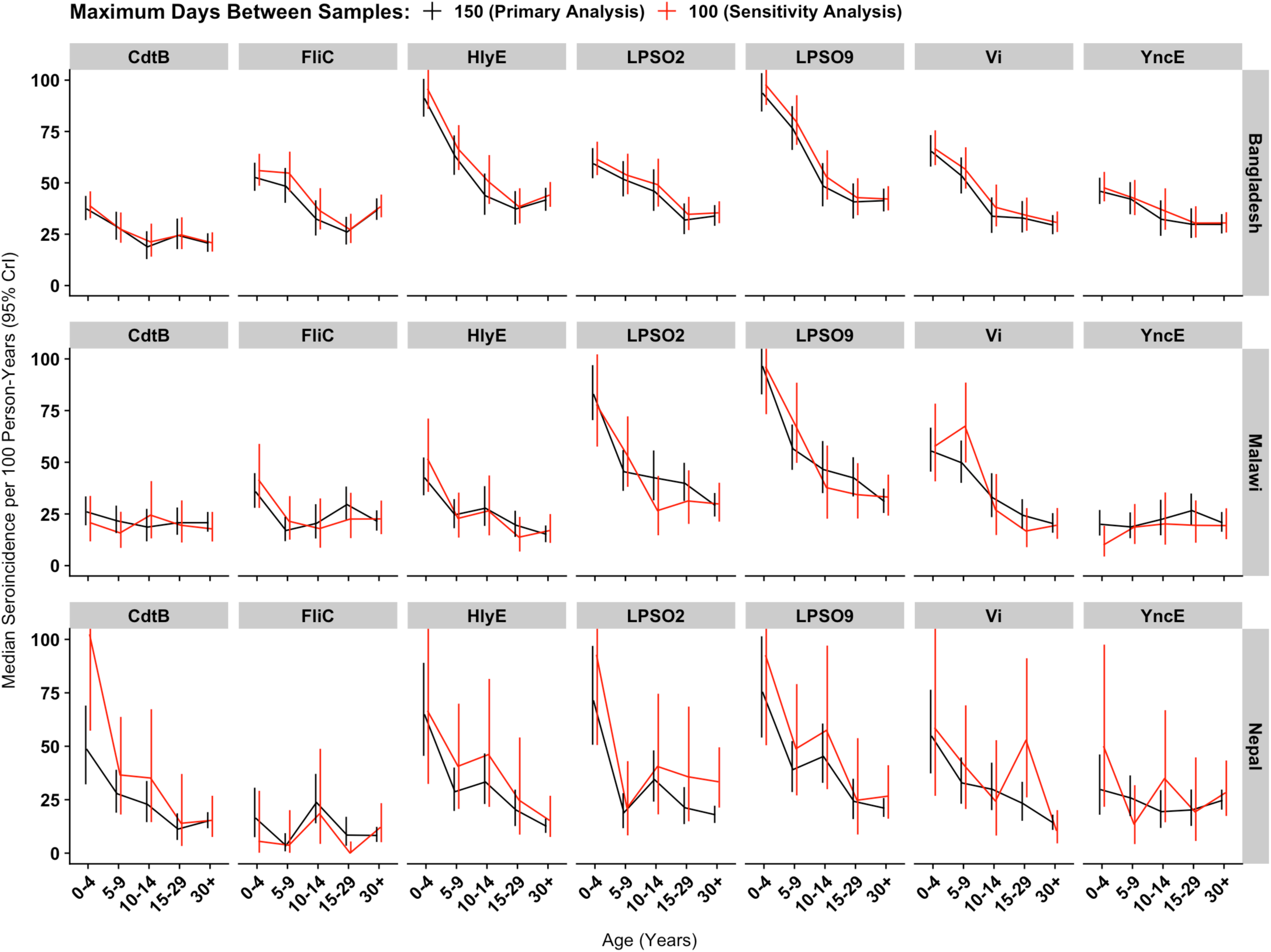
Seroincidence by Maximum Time Between Samples. Each subfigure corresponds to the specific antigen target which was used to classify participants’ infection status when calculating seroincidence. Seroincidence estimates for the Bangladesh, Malawi, and Nepal study sites appear in the top, middle, and bottom row of subfigures, respectively. Solid lines denote the median seroincidence (y-axis) in each age group (x-axis). Vertical lines represent the 95% credible intervals of the seroincidence estimates. Black lines correspond to the primary analysis, which excluded participants with sample pairs collected over 150 days apart, while red lines correspond to a sensitivity analysis which only included participants with < 100 days between samples.

**Supplementary Figure 8:**
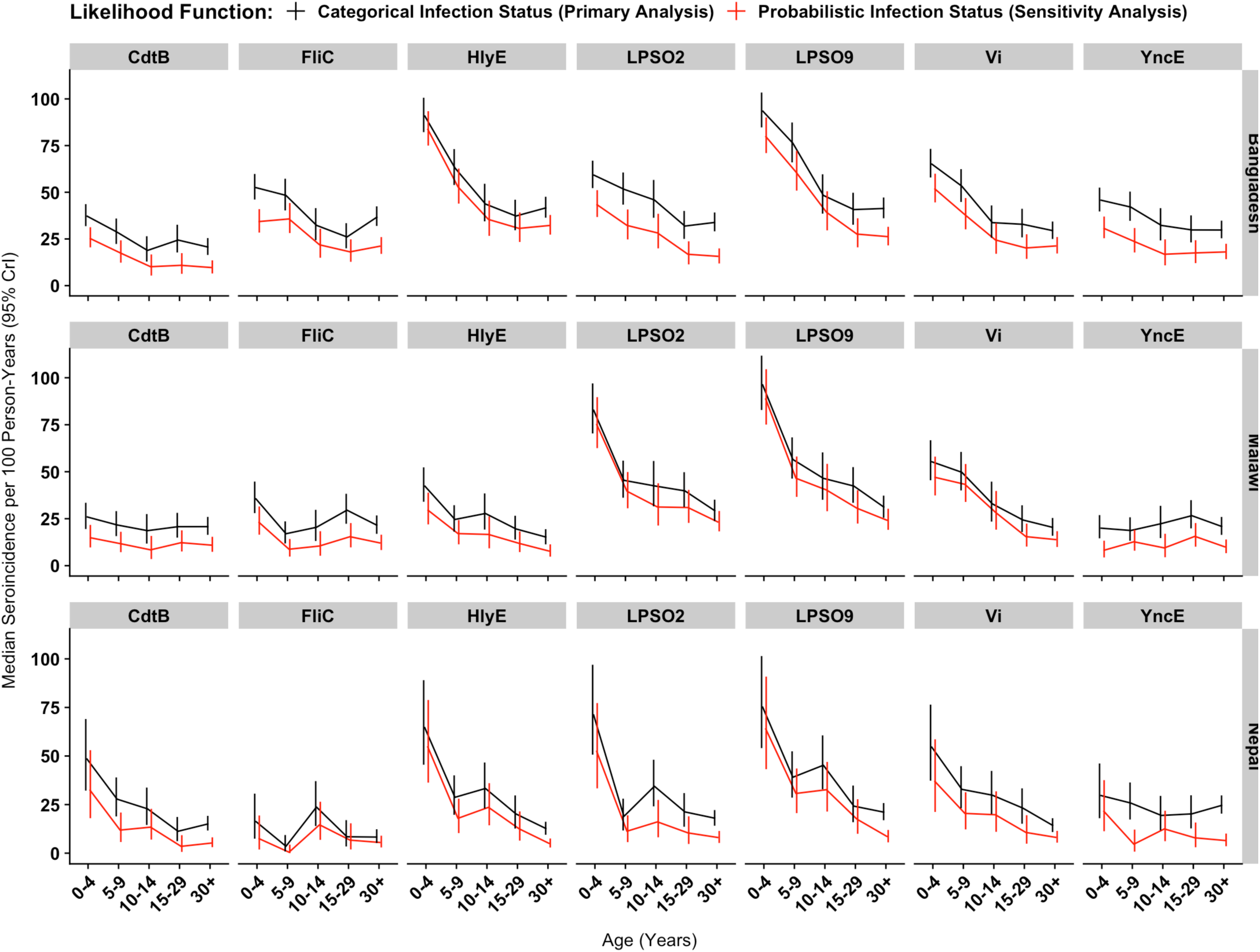
Seroincidence by Likelihood Function. Each subfigure corresponds to the specific antigen target which was used to classify participants’ infection status when calculating seroincidence. Seroincidence estimates for the Bangladesh, Malawi, and Nepal study sites appear in the top, middle, and bottom row of subfigures, respectively. Solid lines denote the median seroincidence (y-axis) in each age group (x-axis). Vertical lines represent the 95% credible intervals of the seroincidence estimates. Black lines correspond to the primary analysis, while red lines correspond to a sensitivity analysis in which the likelihood function for seroincidence directly incorporates participant’s mixture model-derived probability of infection.

**Supplementary Figure 9:**
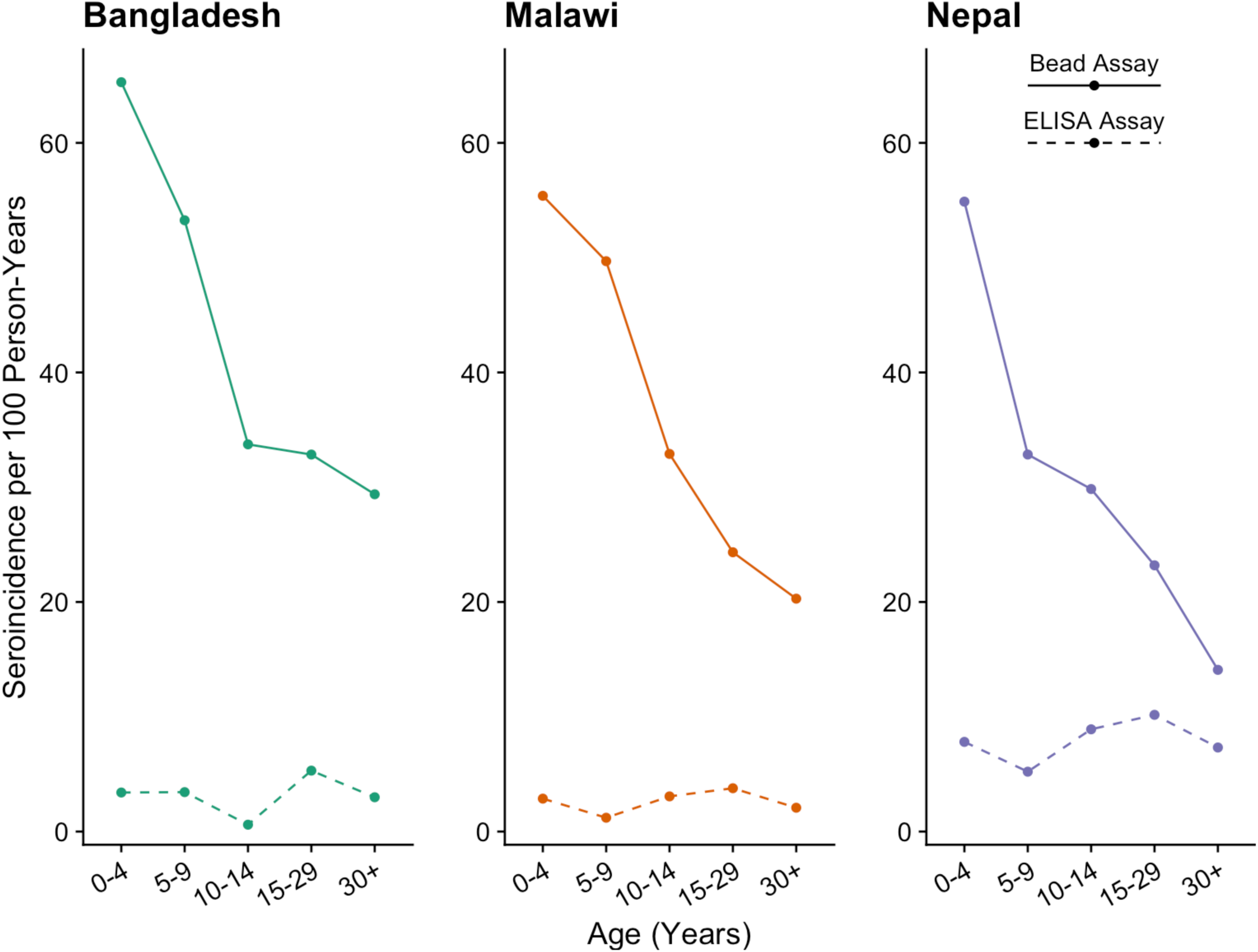
Vi Seroincidence by Age, Study Site, and Assay. Each subfigure corresponds to a different study site. Lines denote the estimated seroincidence (y-axis) in each age group (x-axis). Solid lines represent seroincidence estimates based on the serologic data generated in this study with a bead-based multiplex assay, while the dashed lines depict previously published seroincidence estimates based on ELISA assays. Green, orange, and purple lines correspond to the Bangladesh, Malawi, and Nepal study sites, respectively

